# Genomic and immune determinants of resistance to anti-CD38 monoclonal antibody-based therapy in relapsed refractory multiple myeloma

**DOI:** 10.1101/2023.12.04.23299287

**Authors:** Bachisio Ziccheddu, Claudia Giannotta, Mattia D’Agostino, Giuseppe Bertuglia, Elona Saraci, Stefania Oliva, Elisa Genuardi, Marios Papadimitriou, Benjamin Diamond, Paolo Corradini, David Coffey, Ola Landgren, Niccolò Bolli, Benedetto Bruno, Mario Boccadoro, Massimo Massaia, Francesco Maura, Alessandra Larocca

## Abstract

Anti-CD38 antibody therapies have transformed multiple myeloma (MM) treatment. However, a large fraction of patients inevitably relapses. To understand this, we investigated 32 relapsed MM patients treated with daratumumab, lenalidomide, and dexamethasone (Dara-Rd; NCT03848676). Whole genome sequencing (WGS) before and after treatment pinpointed genomic drivers associated with early progression, including *RPL5* loss and APOBEC mutagenesis. Flow cytometry on 202 blood samples, collected every three months until progression for 31 patients, revealed distinct immune changes significantly impacting clinical outcomes. Progressing patients exhibited significant depletion of CD38+ NK cells, persistence of T cell exhaustion, and reduced depletion of T-reg cells over time. These findings underscore the influence of immune composition and daratumumab-induced immune changes in promoting MM resistance. Integrating genomics and flow cytometry unveiled associations between adverse genomic features and immune patterns. Overall, this study sheds light on the intricate interplay between genomic complexity and the immune microenvironment driving resistance to Dara-Rd.

## INTRODUCTION

In the past two decades, the treatment landscape form multiple myeloma (MM) has undergone remarkable transformation thanks to the development of several novel and highly effective treatments. Among these advancements, the incorporation of targeted immunotherapies, such as anti-CD38 monoclonal antibody (MoAb) daratumumab, stands out as one of the major breakthroughs (Shah and Mailankody, 2020; van de Donk and Usmani, 2018). This development has resulted in significant enhancements in both the depth of treatment response and clinical outcomes, benefiting both newly diagnosed (NDMM) and relapsed/refractory MM (RRMM) patients (Costa et al., 2021; Dimopoulos et al., 2021; Facon et al., 2019; Landgren et al., 2021; Mateos et al., 2018; Rajkumar, 2022; Voorhees et al., 2020). However, despite these improvements, most of the patients inevitably experience relapse and the mechanisms behind this clinical behavior are largely unknown.

Daratumumab works by binding to CD38 on the surface of malignant plasma cells, inducing direct cytotoxic effects and also activating immune-mediated mechanisms like antibody-dependent cellular cytotoxicity (ADCC), complement-dependent cytotoxicity (CDC), antibody-dependent cellular phagocytosis (ADCP), apoptosis induced by cross-linking of CD38 on the target cells, and immunomodulatory effects via elimination of CD38 positive (pos) immunosuppressive cells in the immune tumor microenvironment (TME) (Krejcik et al., 2016; Saltarella et al., 2020; van de Donk et al., 2016). Given the efficacy of anti-CD38 antibody therapies against both tumor and immune cells, particularly when used in combination with immunomodulatory agents (IMIDs) and proteasome inhibitors (PIs), there is a hypothesis suggesting that the resistance of MM to these therapies may stem from a complex interplay between genomic factors within tumor cells and specific patterns within the immune TME. Ideally, this interplay could be investigated by integrating comprehensive genomic and immune data from MM patients treated with daratumumab. However, only a few studies have delved into the mechanisms of treatment resistance to anti-CD38 monoclonal antibodies in MM, with most focusing on either immune profiling of the TME or tumor biology using RNA/FISH-based data (Cohen et al., 2021; Costa et al., 2023; Saltarella et al., 2020; van de Donk and Usmani, 2018; Viola et al., 2021). To comprehensively explore the interplay between the two compartments and track their changes over time, we performed a longitudinal collection of bone marrow (BM) and peripheral blood (PB) samples from a cohort of 32 RRMM patients undergoing treatment with daratumumab, lenalidomide, and dexamethasone (Dara-Rd, ClinicalTrials.gov identifier NCT03848676). To elucidate the genomic factors influencing treatment response or resistance, we conducted whole-genome sequencing (WGS) on CD138+ plasma cells at treatment initiation and disease progression. Concurrently, we employed multi-parametric flow cytometry to characterize the immune TME composition in both PB and BM samples from all 32 patients, prior to treatment initiation. Additionally, we investigated immune alterations over time by analyzing 202 PB samples collected every three months from the onset of treatment until disease progression for each patient (**Figure 1a**). The findings of our comprehensive analysis highlight that resistance to Dara-Rd in RRMM patients stems from an interplay that involves distinct patterns of genomic complexity associated with specific immune profiles. Furthermore, it encompasses specific longitudinal changes in the immune cell composition of the TME. Notably, these changes include the depletion of CD38 pos natural killer (NK) cells, the persistence of CD38 negative (neg) regulatory T-cells (T-reg), and the endurance of exhausted T-cells.

**Figure 1.**
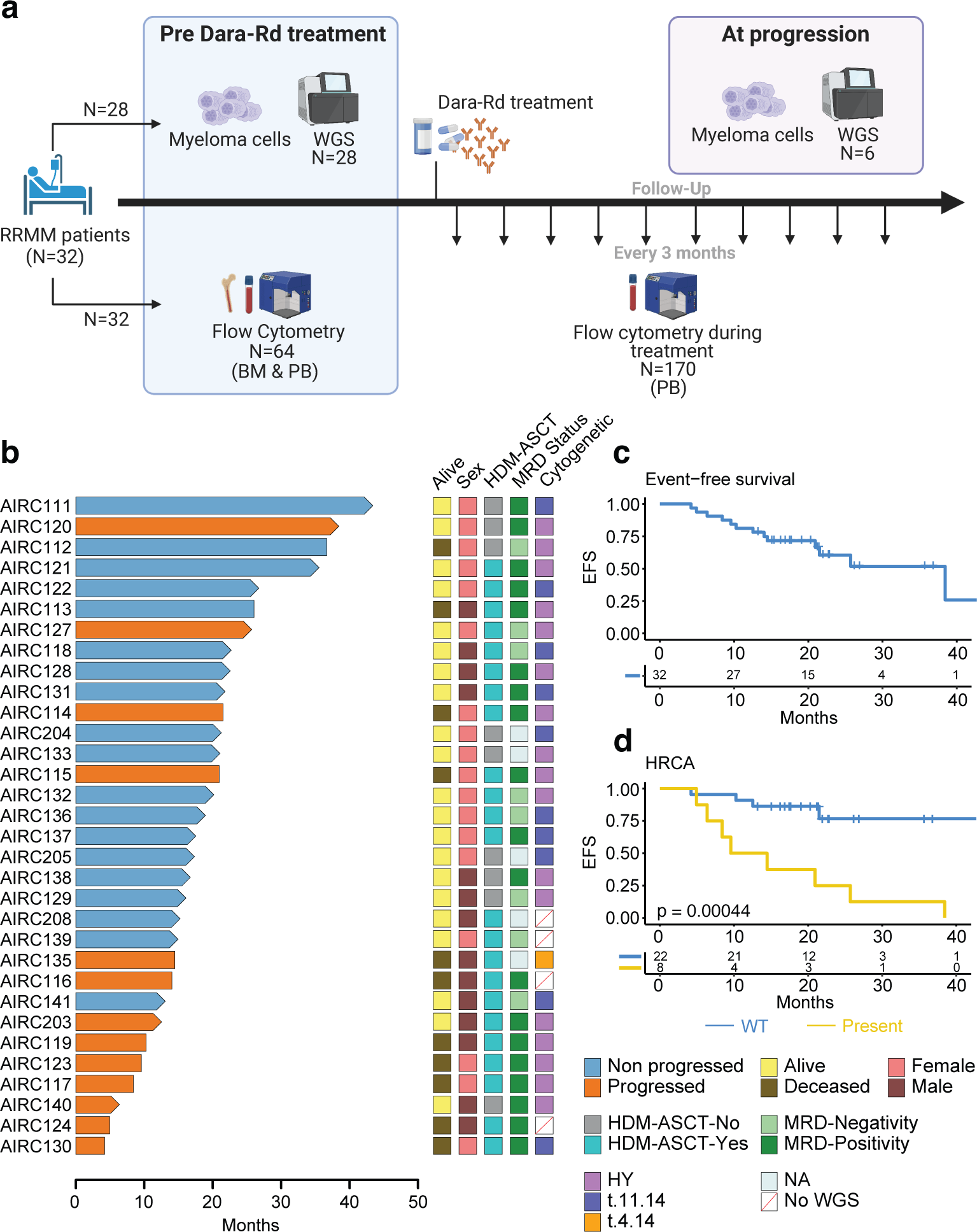
Study design and outcome in RRMM treated with anti-CD38 MoAb. **a)** Study design. **b)** Swimmer plot summarizing the clinical outcome of 32 patients with RRMM enrolled in this study. Arrows indicate patients that were still alive at the last follow up; the no progressed patients (i.e., durable responders) are in light blue, while the progressors in orange. **c)** Even free survival (EFS) Kaplan-Meier curve for the entire cohort. **d)** EFS Kaplan-Meier curve comparing patients with and without the high-risk cytogenetic alterations [HRCA; t(4;14), t(14;16), del17p]; p-values for **c** and **d** was generated with log-rank test. HY: hyperdiploid.

## RESULTS

### Patient cohort

To delineate the genomic and immune factors contributing to resistance against Dara-Rd, we analyzed data from WGS and flow cytometry in a longitudinal cohort of 32 RRMM patients enrolled in the NCT03848676 observational study. Clonal evolution analysis also included two patients, treated with daratumumab single agent and daratumumab, bortezomib and dexamethasone (Dara-Vd). The median age was 68 (range 46-80); 18 (56.2%) were women. The median number of prior treatments was 1 (range 1-3), with 23 (71.8%) patients previously exposed to high-dose melphalan chemotherapy followed by autologous stem cell transplantation (HDM-ASCT; **Figure 1b**). Before starting Dara-Rd, 97% (N=31) and 50% (N=16) of patients were previously exposed to PI and IMIDs, respectively. After a median follow-up of 18.2 months (range 4.2-43.4), 40.6% (N=13) patients experienced progression (i.e., progressors). Patients that did not experience progression were defined as “durable responders”. Overall, 8 (25%) patients achieved minimal residual disease (MRD) negativity by next generation flow cytometry (**Figure 1b**) (Flores-Montero et al., 2017). The median event free survival (EFS) was 38.4 months (**Figure 1c**). Two patients (AIRC112 and AIRC113) died due pulmonary sarcoma and COVID pneumonia without relapse. Considering all the relapsed patients 9 out of 13 (69.2%) died due disease progression. Combining WGS and FISH data, presence of at least one high-risk cytogenetic alterations (HRCA) [i.e., del17p and/or t(4;14) and/or t(14;16)] was observed in 8 out of 30 patients (26.7%; FISH and WGS data was missing in 2 patients) and it was associated with poor EFS (p=0.00044 estimated using Log-rank test; **Figure 1d** and **Supplemental Tables 1-2**) (Costa et al., 2023). We did not see an influence of other key clinical and serological features on the clinical outcome (**Supplemental Table 2**). Demographics, disease characteristics, and response to Dara-Rd for all patients are summarized in **Table 1** and **Supplemental Table 1**.

### Genomic determinant of resistance and progression to Dara-Rd

To investigate the intrinsic genomic drivers within the tumor that confer resistance to Dara-Rd, we performed WGS to profile the genomic landscape in BM samples at baseline and at progression. Four samples out of 32 failed the sequencing due to low cellularity or cancer cell fraction and therefore were excluded. The median coverage for the 28 cases that successfully completed WGS was 70x (**Supplemental Table 3**). The median single base substitution (SBS) burden was 5800 (range 1808-17411). There was no difference in total mutational burden between progressors and durable responders. To assess the clinical implications of major somatic drivers in MM, we utilized an extensive catalog of established myeloma genomic drivers derived from large cohorts of newly diagnosed MM (NDMM) patients (Maura et al., 2022b). that encompasses mutations, mutational signatures, recurrent aneuploidies, and canonical translocations (**Methods** and **Supplemental Table 4**). Among the catalogues of 90 nonsynonymous MM driver mutations (Maura et al., 2019a; Maura et al., 2022b; Walker et al., 2018), we found 31 mutated genes in the RRMM patients treated with Dara-Rd (**Supplemental Figure 1a**). Genes in MAPK pathway (*NRAS*, *KRAS* and *BRAF*) were the most frequently mutated, with 61% of RRMM cases (treated with Dara-Rd) carrying at least one mutation in one of these genes (**Supplemental Figure 1a**). Nevertheless, neither MAPK pathway nor any other mutations in MM driver genes predicted poor anti-CD38 MoAb outcomes (**Supplemental Table 5**). Finally, we did not find any nonsynonymous mutation involving *CD38* gene either before or after treatment.

To investigate underlying mutational processes involved in these cases, we performed a mutational signature analysis (**Methods**) (Alexandrov et al., 2020; Maura et al., 2019b). We identified seven SBS signatures involved in our cohort of RRMM: the clock-like aging signatures (SBS1 and SBS5), APOBEC mutational activity (SBS2 and SBS13), germline center associated polymerase eta somatic hypermutations (SBS9), radical oxygen damage (SBS18), SBS8 and SBS-MM1 (i.e., SBS99; melphalan mutagenesis; **Figure 2a** and **Supplemental Table 6**) (Maura et al., 2019a; Maura et al., 2019b; Maura et al., 2017; Maura et al., 2021; Rustad et al., 2021; Rustad et al., 2020a; Samur et al., 2020; Walker et al., 2015; Weinhold et al., 2016). SBS-MM1 mutational signature was identified in 14 out of 19 (74%) patients who received HDM-ASCT. The lack of SBS-MM1/SBS99 in 5 patients can be explained through the single cell expansion model as previously described (Landau et al., 2020). Specifically, it has been shown that distinct chemotherapy agents promote their mutational activity by introducing a unique catalog of mutations in each exposed single cell (Landau et al., 2020; Pich et al., 2019; Rustad et al., 2020a). These mutations can be detected by bulk WGS only if a single tumor cell exposed to chemotherapy expands, taking clonal dominance. In contrast, if the cancer progression is driven by multiple clones originating from different single cells exposed to chemotherapy, the chemotherapy-induced mutational signature will not be detectable because each clone harbors different catalogs of unique chemotherapy-related variants. Interestingly, two patients (AIRC119 and AIRC128) exhibited two distinct set of melphalan-related variants detected in both subclonal and clonal variants. This scenario was explained by the patients’ exposure to tandem HDM-ASCT. Among the mutational signatures identified in our cohort, high APOBEC contribution was associated to higher rate of progression (p=0.03 estimated using Fisher exact test) and short EFS (p=0.047 estimated using Log-rank test; **Figure 2b** and **Supplemental Table 6**). APOBEC has been shown to be one of the strongest prognostic markers for poor outcomes in NDMM treated with and without daratumumab-based regimens (Maura et al., 2022a; Maura et al., 2017; Maura et al., 2022b; Walker et al., 2015). These findings indicate that the Dara-Rd treatment is not able to entirely overturn the negative prognostic impact of high APOBEC mutational activity. Among the other SBS signatures, the presence of SBS18, known to be caused by reactive oxygen species damage and associated with resistance to CART in diffuse large B-cell lymphoma (Jain et al., 2022), was associated with shorter EFS, with 4 out of 5 patients (80%) progressing after anti-CD38 MoAb combination treatment (p=0.03 estimated using Log-rank test; **Figure 2c** and **Supplemental Table 6**). Finally, high SBS9 showed favorable outcomes, most likely due to the known inverse correlation between APOBEC and SBS9 (p=0.041 estimated using log-rank test; **Supplemental Table 6**) (Maura et al., 2022a; Rustad et al., 2020a).

**Figure 2.**
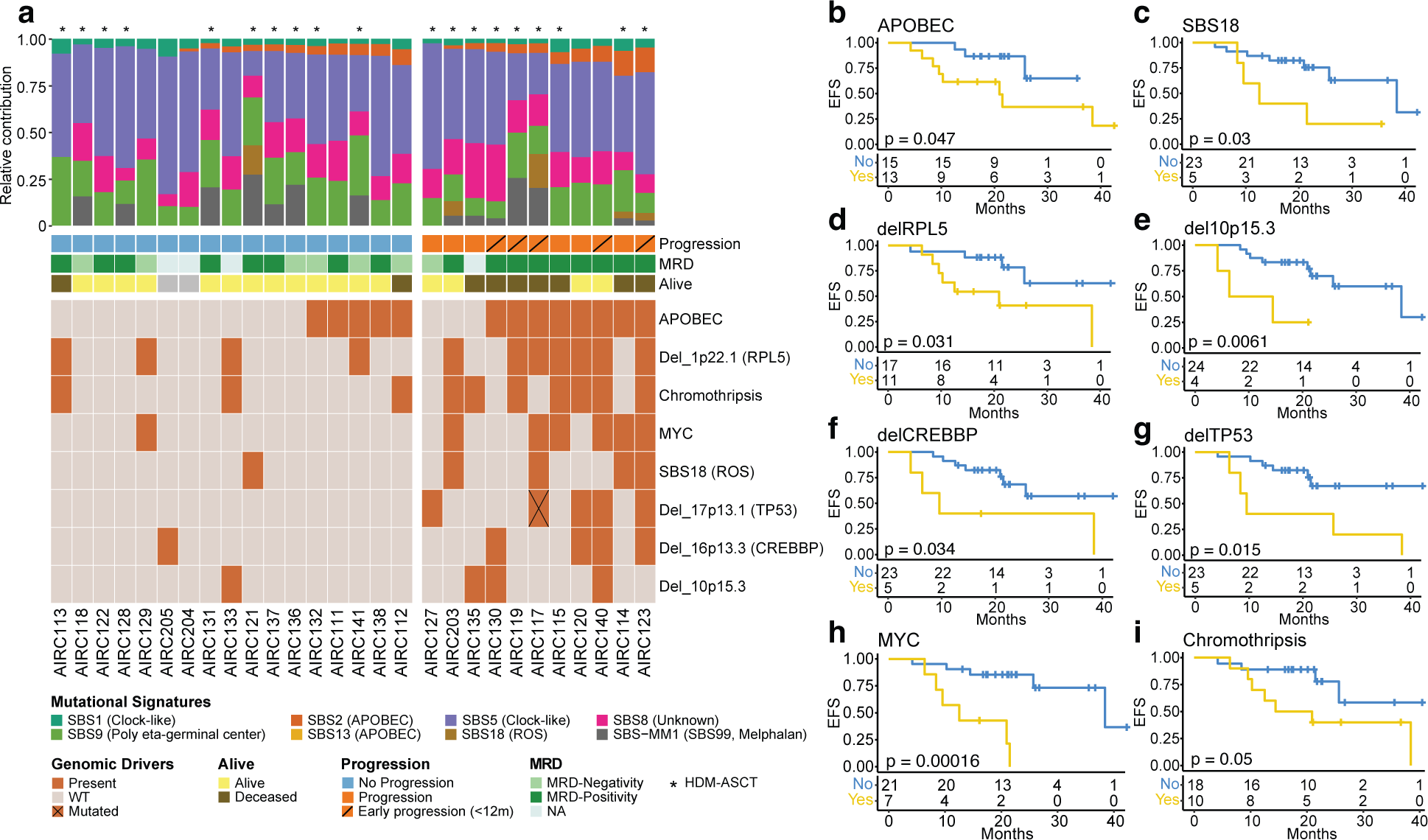
Impact of genomic alterations on clinical outcome in RRMM treated with Dara-Rd. **a)** A heatmap showing all the genomic alterations associated with progression after Dara-Rd among patients with available WGS data (N=28); at the top, the bar-plot with the relative contribution per each SBS mutational signature. Minimal residual disease (MRD) was tested using Euroflow (Flores-Montero et al., 2017). WT: wild type; HDM-ASCT: high-dose melphalan and autologous stem cell transplant; ROS: radical oxygen stress. **b-i)** Kaplan-Meier curves showing the impact of APOBEC (**b**), SBS18 (**c**), deletion 1p22.1 (*RPL5;* **d**), deletion 10p15.3 (**e**), deletion 16p13.3 (*CREBBP*; **f**), deletion 17p13.1 (*TP53*; **g**), structural variants on *MYC* (*MYC*, *PTV1* and *NSMCE2*; **h**) and presence of chromothripsis (**i**) on event free survival (EFS); p-value is calculated with log-rank test.

Next we went to investigate the clinical impact of recurrent MM aneuploidies and structural variants (**Supplemental Figure 1b**, **Methods**) (Maura et al., 2022b). Different deletions were associated with shorter EFS. Specifically, deletion of 1p22.1 (*RPL5*) was associated with shorter EFS (p=0.031 estimated using Log-rank test; **Figure 2d**). Mono and bi-allelic loss of *RPL5* have been associated with poor response in both Dara and non-Dara-based regimens (Hofman et al., 2017; Maura et al., 2022a). In addition to *RPL5*, patients with loss of 10p15.3, 16p13.3 (*CREBBP*) and *TP53* were characterized by shorter EFS (p=0.0061, p=0.034 and p=0.015 estimated using Log-rank test, respectively; **Figure 2e-g**). Since the identified aneuploidies 16p13.3 and 10p15.3 are genomic event not extensively characterized in the literature, we investigated their biological impact using paired WGS and RNA sequencing data from 705 patient enrolled in the CoMMpass study (https://themmrf.org). Running a gene set enrichment analysis (GSEA), we found that patients harboring one of these two alterations (versus wild-type) were enriched for *E2F, MYC* targets and *G2M* checkpoint pathways (**Supplemental Figure 2**). These pathways were also found to be enriched in RRMM patients that progressed after Daratumumab, Carfilzomib, Lenalidomide and dexamethasone at first relapse (Kydar study) (Cohen et al., 2021; Maura et al., 2022a). These data suggest that 16p13.3 and 10p15.3 deletions can contribute to the deregulation of distinct high-risk pathways associated with resistance to Dara-Rd treatment.

Thanks to WGS resolution we were able to investigate the clinical impact of structural variants (SVs) and complex events in patients treated with Dara-Rd (**Supplemental Figure 1b**) (Rustad et al., 2020b). SVs causing *MYC* gain of function were associated with shorter EFS, similarly to what observed in NDMM treated with Dara-KRd (i.e., *NSMCE2* deletions, *MYC* and *PVT1* events; p=0.00016 estimated using Log-rank test; **Figure 2h**) (Maura et al., 2022a). Among complex events, presence of chromothripsis, a well-recognized adverse prognostic factor marker in NDMM, was found in 36% (10/28) of patients, with 7 experiencing progression (p=0.05 estimated using Log-rank test; **Figure 2i**) (Rustad et al., 2020b). Genomic events associated with shorted EFS were not differentially distributed between early (EFS<12 months) and late progressors. Overall, these data suggest that distinct genomic events and patterns of genomic complexity are associated with resistance to Dara-Rd treatment.

### Clonal evolution post Dara-Rd treatment

To reconstruct the genomic evolution from baseline to progression after Dara-based treatment, we analyzed WGS data from four patients with longitudinal paired samples collected before treatment and at progression. Additionally, we included two patients treated with daratumumab as a single agent or in combination with bortezomib and dexamethasone (Dara-Vd), with available WGS data before and after treatment. Within these six patients, we identified patterns of branching evolution in all cases, with 50% experiencing a complete clonal shift (**Figure 3a-f**). The presence of clonal shift was independent of the patients’ EFS. Although we didn’t observe any recurrent genomic events, we did detect distinct and intriguing events that were undetectable at baseline but became dominant upon progression. In the patient AIRC104 the progression was driven by a clone with a previously undetectable chromothripsis event involving chromosome 4, but not *CD38* (**Figure 3a**). In AIRC114 and AIRC115 the progression was driven by clone carrying translocations between chromosome 4 and 17 involving *IRF4* and *IKZF3* genes, and a large deletion of *TP53*, respectively (**Figure 3c-d**). In AIRC117 at progression the dominant clone had a deletion of chromosome 4p involving *CD38* undetectable at baseline (**Figure 3e**).

**Figure 3.**
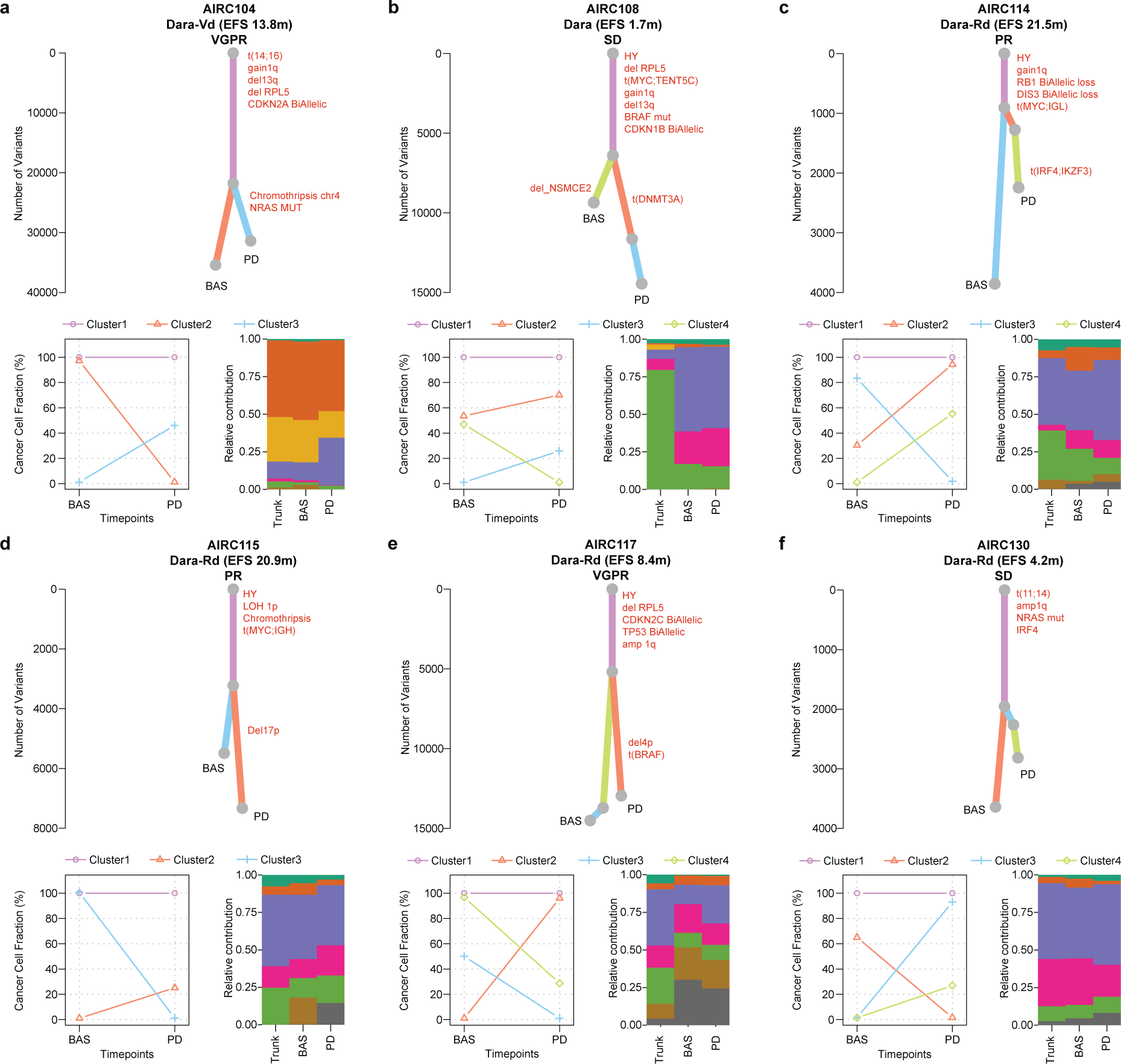
Clonal evolution of RRMM patients treated with daratumumab based treatment. Top of each panel: the phylogenetic tree with the trunk in violet and the branches in salmon, light blue, and light green. The y-axis is the number of single nucleotide variants. At the bottom left side, the cancer cell fraction (CCF) scatter-plot per each cluster at baseline (BAS) and progression (PD). In the bottom right side, the bar-plot with the mutational signature relative contribution for each cluster. The mutational signature color legend is the same of the Figure 2. HY: hyperdiploid.

### Association between immune profiles at baseline and clinical outcomes

To investigate the immune composition and its impact on clinical outcomes, BM and PB samples collected at baseline from 32 patients were investigated by flow cytometry (see **Methods; Supplemental Table 7**). In line with what previously shown, both the relative and absolute number of immune cells were highly correlated between BM and PB, with 77% of cell types having Spearman coefficient equal or greater than 0.50 (p<0.001; **Supplemental Figure 3** and **Supplemental Table 8**) (Coffey et al., 2023; Zavidij et al., 2020). Next, we went to investigate the association between distinct immune patterns and MM response to Dara-Rd. First, we characterized the immune cell landscape with particular attention to CD38 pos immune cells and pattern of immune exhaustion. This analysis revealed that most myeloid-derived suppressor cells (MDSC), monocytes, and natural killer (NK) cells were CD38 pos in both BM and PB samples (**Supplemental Figure 4a**). Noticeably lower counts and proportions of CD38 pos cells were observed in conventional helper and cytotoxic T-cells, Vψ9Vο2 T-cells, B-reg and T-reg (**Supplemental Figure 4a**). Despite monocytic-MDSCs (Mo-MDSCs) exhibited a higher CD38 pos expression compared to granulocytic-MDSCs (Gr-MDSCs) at the baseline in both BM and PB (**Supplemental Figure 4b**), they were less prevalent than Gr-MDSCs (**Supplemental Figure 4c**). Furthermore, at baseline all patients showed high prevalence of CD38 pos classical and intermediate monocytes, and the classical subgroup was the most predominant compared with intermediate and non-classical monocytes (**Supplemental Figure 4d-e**). In addition, at baseline in BM samples the fraction of CD38 pos was higher among cytotoxic NK cells than proliferative NK cells (**Supplemental Figure 4f**). Across the whole BM immune populations, the proportion of cytotoxic NK cells was less abundant than the proliferative NK cells (**Supplemental Figure 4g**).

Next, we went to investigate the impact of the BM and PB immune composition on clinical outcomes. At baseline patients who experienced progression after Dara-Rd showed an enrichment of CD38 pos NK cells (p=0.04 in the BM and p=0.05 in the PB estimated by Wilcoxon test; **Figure 4a** and **Supplemental Figure 5a-b**), and CD38 pos Mo-MDSC and Gr-MDSC cells compared to the durable responders (p=0.033 and p=0.01 estimated by Wilcoxon test, respectively in the BM only; **Supplemental Figure 5a**). Interestingly, at baseline, patients who progressed were enriched in higher levels of exhausted T-cells, such as TIM3+ cytotoxic T-cells and TIM3+ helper T-cells (p=0.0089 and p=0.0031 respectively; **Supplemental Figure 5c-f**) in PB, and lower presence of helper T-cells (p=0.015; **Supplemental Figure 5g-h**) in BM. Moreover, at baseline a significant enrichment of classical and intermediate monocytes was identified in progressed patient PB samples (p=0.039 both; **Supplemental Figure 5i-l**). These data suggest that Dara-Rd efficacy in RRMM patients is influenced by the presence of distinct patterns of immune composition at baseline, in particular high CD38 pos immune cells, exhausted T-cells, and reduced helper T-cells in the BM and/or PB.

**Figure 4.**
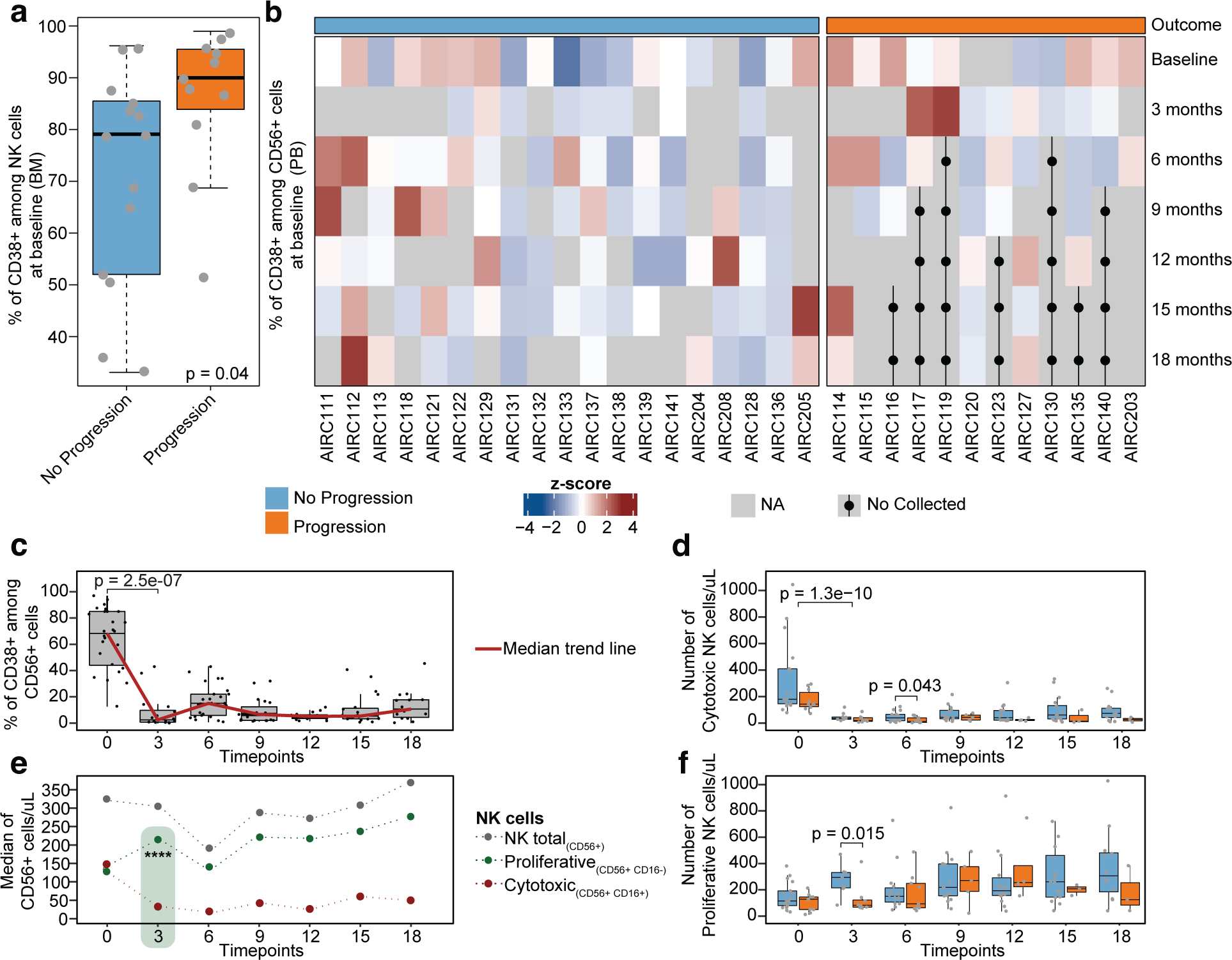
Daratumumab immunomodulation on Natural Killer cells. **a)** Boxplot showing the percentage of CD38 pos among NK cells (CD56 pos). Light blue indicates durable responders, orange the progressors. p-value was calculated with Wilcoxon test two sided. **b)** Heatmap showing CD38 pos in CD56 pos NK cells z-scores per each patient at 7 different timepoints. Dots and black lines indicate that the sample was not collected at that time point, as the patient has previously progressed; one patient (AIRC124) was excluded because several samples were not collected at different time points. **c)** Boxplots displaying the percentage of CD38 pos among CD56 pos NK cells overtime. The red line connects the median value at each time point. **d)** Boxplots showing the absolute number of cytotoxic NK cells overtime. **e)** Smooth plot presenting different NK cells patterns over time; ****≤0.0001. **f)** Boxplots showing the number of proliferative NK cells overtime. In **c-f)** the p-values were estimated using Wilcoxon test two sided. The full list of all p-values is reported in the **Supplemental Table 10-11**.

### Immune modulation induced by Dara-Rd treatment over time

Given the concordance observed between BM and PB samples, we conducted a longitudinal investigation to assess how Dara-Rd treatment influenced the immune system’s composition over time. This investigation involved the analysis of 202 PB samples collected at various time points from 31 patients enrolled in this study. (**Supplemental Table 7**). One sample was excluded (AIRC124) due to lack of longitudinal samples after Dara-Rd treatment. As expected, both CD38 pos tumor and normal cells were rapidly depleted after exposure to daratumumab (Casneuf et al., 2017; Maura et al., 2022a; Viola et al., 2021) (**Supplemental Figure 6a** and **Supplemental Table 9**). Among the main immune cell populations, a significant depletion was observed among cytotoxic and CD38 pos NK cells immediately after Dara-Rd treatment (p<0.0001 estimated by Wilcoxon test at 3 month; **Figure 4b-d**). In contrast, we observed a significant expansion of proliferative NK cells at 3 months (p<0.0001 estimated by Wilcoxon test; **Figure 4e** and **Supplemental Table 10**). This expansion was driven by CD38 neg proliferative NK cells that were not depleted by the daratumumab. Interestingly, the expansion of proliferative NK cells was significantly higher in durable responders compared to progressors (p=0.015 estimated by Wilcoxon test; **Figure 4f** and **Supplemental Table 11**). Durable responders were also characterized by a significant expansion of cytotoxic NK cells at 6 months (p=0.043 estimated by Wilcoxon test, **Figure 4d** and **Supplemental Table 11**). Consistent with earlier research, this data underscores the critical role of NK cells in the efficacy of anti-CD38 MoAb-based treatments (Casneuf et al., 2017; Maura et al., 2022a; Viola et al., 2021). Furthermore, these findings suggest a scenario in which daratumumab facilitates the fratricide depletion of CD38 pos NK cells, potentially constraining the subsequent availability of immune effector cells in the fight against myeloma cells.

Next, we investigated the immune dynamics of monocytes cells and the classical (CD14++ CD16-), intermediate (CD14++ CD16+) and non-classical (CD14+ CD16++) subgroups. The longitudinal analysis showed a significant reduction of CD38 pos monocytes at 3 months after Dara-Rd treatment (p=0.0001 estimated by Wilcoxon test), and the reduction was particularly relevant among the classical and intermediate subgroups (p=0.0005 and p=0.03 estimated by Wilcoxon test, respectively; **Supplemental Figure 6b** and **Supplemental Table 10**). Interestingly, after being depleted at 3 months, the CD38 pos intermediate monocytes experienced a significant expansion after 12 months of therapy (p=0.0042 estimated by Wilcoxon test; **Supplemental Figure 6b** and **Supplemental Table 10**). In addition, independently from the CD38 status, reduction of intermediate and non-classical monocyte subgroups was observed at 3 months after Dara-Rd treatment (p=0.0036 and p=0.0001 estimated by Wilcoxon test; **Supplemental Figure 6c-d** and **Supplemental Table 10**).

Finally, we investigated the composition and clinical implications of T-reg and of patterns of T-cell exhaustion. While the proportion of T-reg (i.e., CD4+ CD25++ FOXP3+) over both the total number of immune cells and the T-CD4 experienced a significant expansion over time, in particular after 9 months across all patients (**Figure 5a** and **Supplemental Table 10**), the CD38 pos T-reg experienced a significant and sustained depletion immediately after the first cycles of Dara-Rd (p<0.0001 estimated by Wilcoxon test, **Figure 5a**, **Supplemental Figure 6e**, and **Supplemental Table 10**). While the proportion of CD38 pos T-reg was not significantly different between durable responders and progressors, the latter group had a higher proportion of all T-reg (i.e. CD38 pos and CD38 neg) over the total number of T-CD4 at 3 months compared to durable responders (p=0.05 estimated by Wilcoxon test; **Figure 5b**; **Supplemental Table 11**). In term of T-cell exhaustion, we found an increase in LAG3+ T-cells following Dara-Rd treatment, particularly LAG3+ helper T-cells (p=0.014 estimated by Wilcoxon test; **Figure 5c**; **Supplemental Table 11**). Notably, this increase was observed consistently over time. Furthermore, progressors showed higher expansion of LAG3+ helper T-cells at 15 months compared to durable responders (p=0.013 estimated by Wilcoxon test; **Figure 5c**; **Supplemental Table 11**). Additionally, progressors had a higher proportion of Gr-MDSC at 15 months (p=0.0025 estimated by Wilcoxon test; **Figure 5b**; **Supplemental Table 11**). Conversely, the durable responders showed an enrichment of helper T-cells (p=0.046 at 6 months) and Vψ9Vο2 T-cells at different timepoints (i.e., p=0.048 at 3 months and p=0.008 at 12 months estimated by Wilcoxon test; **Figure 5b**; **Supplemental Table 11**). These findings indicate that progression after Dara-Rd in RRMM patients is associated with distinct changes over time in the immune compositions with immunosuppressed and immune-exhausted patterns associated with progression.

**Figure 5.**
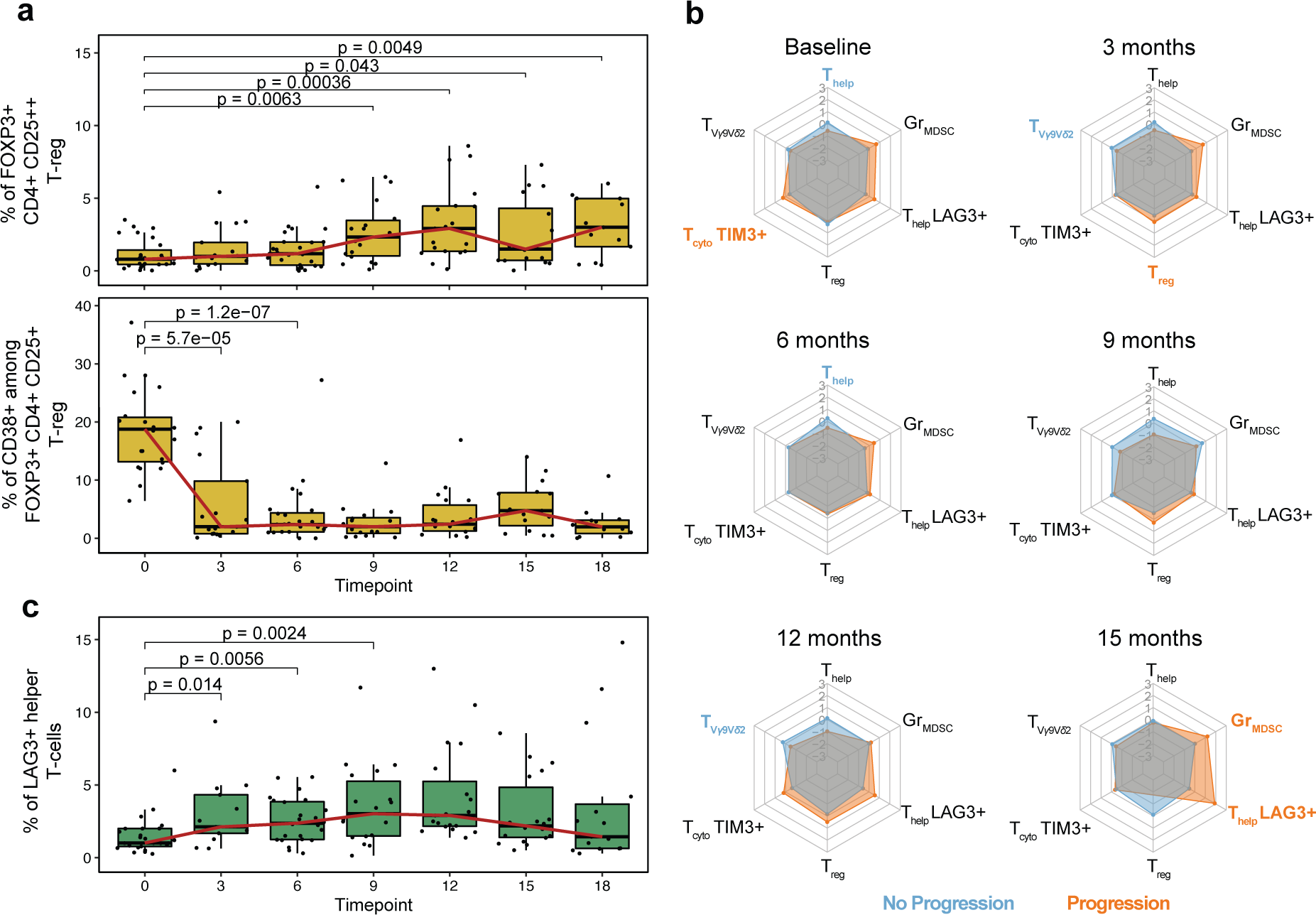
Regulatory and exhausted T-cells changes over time. **a)** Boxplot showing the percentage of FOXP3 T-regs among all T-CD4 (top) and the percentage of CD38 pos among FOXP3 T-regs (bottom). The red line connects the median value at each time point. p-values were estimated using Wilcoxon test two sided. **b)** Radar-plots for each timepoint reporting the contribution of each cell types. The grey network indicates the z-scores values (from -3 to +3), each corner represents an immune cell type; the values represented are the median values of the z-scores for each cell type; the light blue hexagons refer to the values of durable responders, while the orange ones refer to the progressors. Bold and colored cell type names indicate that the specific cell type was significantly enriched at that time point. z-scores were computed using the relative values. T_reg_ = FOXP3+ T-reg. All p-values were estimated using Wilcoxon test two sided. **c)** Boxplot displaying the percentage of LAG3 helper T-cells among all T-CD4 over time. The red line connects the median value at each time point. p-values were estimated using Wilcoxon test two sided. The full list of all p-values is in the **Supplemental Table 10**.

### Interplay between intrinsic-tumor and extrinsic-immune features

Because our findings showed that both genomics drivers and immune patterns can influence clinical outcomes in RRMM treated with Dara-Rd, we integrated WGS and flow cytometry data to define possible link between the two compartments. Integrating significant genomic features and distinct immune cell population in 28 patients at baseline, significant associations were observed (**Figure 6a and Supplemental Table 12**). Specifically, patients with high APOBEC mutational activity were characterized by an enrichment of exhausted TIM3+ helper T-cells (p=0.02; **Figure 6b**), and exhausted cytotoxic T-cell expressing *LAG3* and *TIM3* transmembrane protein (p=0.015 and p=0.048 estimated by Wilcoxon test, respectively; **Figure 6c** and **Supplemental Figure 6f**). Patients without reactive oxygen species damage mutational activity (SBS18) were enriched in cytotoxic NK cells (p=0.044 estimated by Wilcoxon test; **Supplemental Figure 6g**). and CD38 pos monocytes (p=0.036 estimated by Wilcoxon test; **Supplemental Figure 6h**). SVs involving *MYC* were associated with exhausted TIM3+ cytotoxic T-cells (p=0.018 estimated by Wilcoxon test; **Figure 6d**) and have less naïve helper T-cells (p=0.017 estimated by Wilcoxon test; **Supplemental Figure 6i**). Patients with 10p15.3 deletions showed high LAG3+ helper T-cells (p=0.024 estimated by Wilcoxon test; **Supplemental Figure 6l**) and central memory cytotoxic T-cells (p=0.048 estimated by Wilcoxon test; **Supplemental Figure 6m**). In addition, deletions involving 1p22.1 (*RPL5)* and presence of chromothripsis were associated with low CD38 pos T-reg cell levels (p=0.041 estimated by Wilcoxon test; **Figure 6e**) and an expansion of non-classical monocytes in progressed RRMM patients (p=0.015 estimated by Wilcoxon test; **Supplemental Figure 6n**). Overall, reduced baseline number of NK cells was observed in all genomic features associated with progression (p=0.024 estimated by Wilcoxon test; **Figure 6f**). Overall, this integrated analysis revealed an association between distinct genomic and distinct immune patterns, which contributed to promote resistance to Dara-Rd. On the other hand, non-progressed patients show a less impaired genome and a less exhausted and immunosuppressed immune composition.

**Figure 6.**
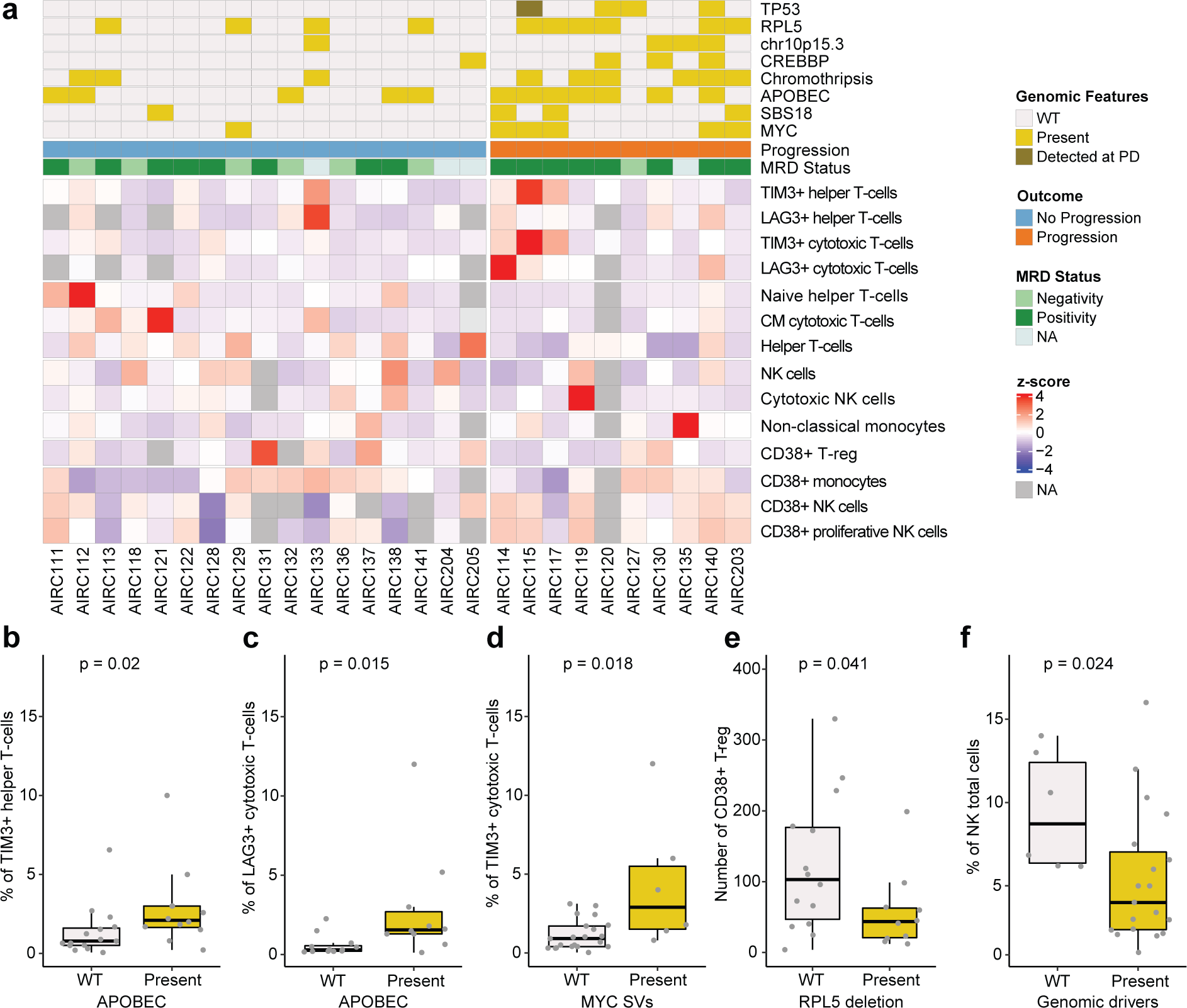
Association between intrinsic tumor drivers and microenvironment cell populations. **a)** Heatmap showing the BM immune cell composition at baseline that were significantly associated with genomic drivers and outcomes. One patient (AIRC123) was excluded because without baseline BM sample. **b-f)** Boxplots showing the distribution of the immune cells according to the presence of absence of distinct genomic drivers linked to shorter event free survival (EFS). The color legend is the same of the panel **a**. All p-values were computed using Wilcoxon test two sided.

## DISCUSSION

The advent of immune active therapies has significantly improved clinical outcomes in RRMM patients (Costa et al., 2021; Dimopoulos et al., 2021; Facon et al., 2019; Landgren et al., 2021; Mateos et al., 2018; Rajkumar, 2022; Voorhees et al., 2020). Despite these improvements it is unknown why a large fraction of patients fails these new therapeutic approaches. To comprehensive characterize the intrinsic and extrinsic mechanisms of resistance to daratumumab-based regimen, we integrated WGS data from the tumor cells and flow cytometry of the BM and PB immune composition in 32 RRMM patients. Thanks to an extensive longitudinal collection of 202 PB samples, we were able to track over time how Dara-Rd affect immune composition and how this was associated with early progression. Our data revealed that RRMM patient progression after Dara-Rd is driven by an intricate interplay between tumor intrinsic genomic features together with changes in the extrinsic immune TME. In line with prior evidence in NDMM patients treated with and without Daratumumab, high APOBEC mutational activity, chromothripsis events, *TP53* deletions and 1p22.1 deletions (*RPL5*), were associated with worse outcomes (Costa et al., 2023; Hofman et al., 2017; Maura et al., 2022a; Maura et al., 2017; Maura et al., 2022b; Rustad et al., 2020b; Walker et al., 2015; Ziccheddu et al., 2020). *RPL5* loss, recently associated with adverse outcomes after Dara-KRd (Maura et al., 2022a) and VRd (Maura et al., 2022b), has been linked to resistance to proteasome inhibitors due to its biological role in protein transport and regulation (Hofman et al., 2017). Interestingly, in this study, *RPL5* was associated with a shorter EFS in patients treated without PIs, suggesting a broader role in promoting resistance to various anti-MM therapies. Interestingly, we found SVs involving *MYC* locus were correlated with early progression. Likewise, *MYC* was recently associated with early progression after Dara-KRd treatment in NDMM patients (Maura et al., 2022a). Moreover, we identified two novel deletions linked to resistance, involving 16p13.3, *CREBBP* tumor suppressor genes, and 10p15.3, both involved in the dysregulation of critical specific pathways, such as *E2F*, *MYC* targets and *G2M* checkpoint pathway that have reported to be enriched in MM that fails daratumumab-based regimens (Cohen et al., 2021; Maura et al., 2022a). Finally, these genomic features, in particular APOBEC, have been linked to distinct microenvironment exhaustion, genomic instability, immune escape and inflammatory profile. In summary, our data demonstrate the influence of genomic complexity on the response to modern immunotherapies, highlighting how WGS can comprehensively characterize this complexity and guide the development of personalized treatment strategies.

Through the application of a longitudinal design, we confirmed and expanded the notion that immune patterns at baseline and their changes over time are strongly associated with response and clinical outcomes in MM receiving daratumumab-based regimens (Casneuf et al., 2017; Maura et al., 2022a; Viola et al., 2021). Overall, we found three key immune patterns. The first involves NK cells and the ADCC daratumumab mechanisms of action. Specifically, patients who progressed showed a higher level of CD38 pos NK cells at baseline, which was rapidly depleted as soon as they received the anti-CD38 MoAb treatment. Moreover, in all patients cytotoxic NK cells (CD56+ CD16+) develops a drastic reduction after daratumumab treatment, while the proliferative NK cells (CD56+ CD16-) expanded after Dara-Rd treatment in the entire cohort (Poli et al., 2009). The increase was primarily driven by CD38 neg proliferative NK cells and was notably higher among durable responders. Moreover, in durable responders, cytotoxic NK cells expanded more at 6 months compared to progressors. This scenario reveals how a less activated immune environment retains a higher pool of NK cells, non-depleted by daratumumab exposure. These NK cells subsequently expand, significantly contributing to the anti-tumor immune activity of daratumumab. Conversely, a more activated immune environment, dominated by CD38 pos NK cells, experiences major NK cells depletion after daratumumab, thereby limiting subsequent anti-tumor activity. The second pattern involves FOXP3+ T-reg cells that expand over time, in particular among progressors. The persistence and expansion of these cells can promote an immunosuppressed environment, affecting the daratumumab-mediated immune activity against MM. The third and final pattern revealed how presence and persistence of exhausted T-cell at baseline associated with worse clinical outcomes, in line with what observed with other immunotherapy agents in RRMM (Dhodapkar et al., 2022; Friedrich et al., 2023).

Overall, the study presented here provides novel insights that contribute to our understanding of resistance to emerging anti-CD38 monoclonal antibodies in MM. Furthermore, it underscores the critical need for integrated multi-omics approaches to thoroughly analyze the tumor cell and immune heterogeneity in MM. This understanding will be instrumental in redefining the concept of high-risk MM and enhancing patient management through the application of personalized medicine.

## METHODS

### Patients

32 relapsed/refractory MM patients were enrolled after informed consent and treated with Dara-Rd therapy (NCT03848676). Summary of key clinical and treatment data are available in **Table 1** and **Supplemental Table 1**.

### Sample processing

CD138-positive and germline cells were collected from bone marrow (BM) and (PB) diagnostic samples, respectively. Briefly, Bone marrow plasma cells were enriched using anti-CD138-coated magnetic microbeads and AutoMACS Pro Separator (Miltenyi Biotec GmbH, Bergisch Gladbach, Germany) as previously described (Oliva S Ann Hematol 2021). Purity was assessed by multiparameter flow cytometry (plasma cell purity always exceeded 90%). Differently, germline cells were separated using Ficoll density stratification followed by red cells lysis procedures. Finally, both samples were stored at −80 °C until genomic DNA extraction.

### DNA extraction

CD138-positive and germline DNA were extracted using commercially available kit Maxwell RSC Blood Kit (Promega, Madison, USA) following manufacturer’s instructions. DNA quantification was performed employing both photometric (NanoDrop/Thermofischer) and fluorescent systems (Quantus; Promega, Madison, USA). 260/280 rand 260/230 ratios were employed to estimate DNA quality.

### Whole genome sequencing

Thirty-two matched tumor-normal samples underwent whole genome sequencing (WGS) at the San Raffaele Institute. In short, libraries were prepared using the Illumina Nextera DNA Flex protocol. Samples were run on NovaSeq 6000 in a 2x150 bp paired end (Illumina).

### Whole genome sequencing analysis

The pre-processing steps and the variant file generation have been performed at San Raffaele Institute. The nf-core/sarek pipeline (https://github.com/nf-core/sarek) was used to align tumor and normal paired samples against the GRCh38 reference genome using the Burrows–Wheeler Aligner. All samples were uniformly analyzed by the following bioinformatic tools: somatic mutations were identified by integrating MuTect2 and Strelka2, selecting the variants called by both callers; copy number analysis and tumor purity (i.e., cancer cell fraction) were evaluated using ASCAT; structural variants were defined by Manta algorithm. Complex events (i.e., templated insertions, chromothripsis, and chromoplexy) were defined after manual inspection, as described previously (Rustad et al., 2020b). To determine the tumor clonal architecture, we used PyClone algorithm (v0.13.1; https://github.com/Roth-Lab/pyclone).

### Mutational signatures

Mutational signatures were analyzed across all whole genomes. To estimate the activity of mutational signatures, we first used a three-step approach of de novo extraction, assignment, and fitting (Maura et al., 2019b; Rustad et al., 2021). For the first step, we ran SigProfiler for SBS signatures. All extracted signatures were then compared with the latest Catalogue of Somatic Mutations in Cancer (https://cancer.sanger.ac.uk/cosmic/signatures/SBS; COSMIC) to identify the known mutational processes active in the cohort. Finally, we applied mmsig (https://github.com/UM-Myeloma-Genomics/mmsig) (Rustad et al., 2021), a fitting algorithm, to confirm the presence and estimate the contribution of each mutational signature in each sample guided by the catalog of signatures extracted for each individual sample adding the chemotherapy SBS-MM1 signature for patients who received melphalan (Maura et al., 2021).

### RNAseq analysis and Gene Set Enrichment analysis (GSEA)

The raw count data was filtered to remove genes with less than 10 reads in greater than 95% of samples. The trimmed mean of M-values (TMM) normalization was applied (Robinson and Oshlack, 2010).This method estimates a scale factor used to reduce technical bias between samples with different library sizes. The voom transformation was performed to convert row counts in log2-counts per million (log2-CPM) and calculated the respective observation-level weights to be used in differential expression analysis (Law et al., 2014). P values were corrected for multiple testing using Benjamini-Hochberg false discovery rate method. The Gen Set Enrichment Analysis (GSEA) was performed using the *fgsea* R package (Subramanian et al., 2005). The H Hallmark gene sets collection, retrieved from MSigDb database v7.4 (Liberzon et al., 2011), was enriched with two INF signatures, ISG.RS and IFNG.GS, previously described to be associated with response to immunotherapy (Jain et al., 2021). Genes were ranked using the statistic derived from differential expression analysis with voom/limma pipeline (Law et al., 2014).

### Analysis of malignant plasma scRNA in the Kydar study

scRNAseq analysis was performed on CD138^+^/CD38^+^ FACS-sorted plasma cells from a previously published Kydar study of 41 patients with MM who failed to respond to a bortezomib-containing induction regimen and received Dara-KRd (Cohen et al., 2021). Gene expression matrices were normalized, scaled, and integrated using Seurat v4 and cells with less than < 500 unique molecular identifiers, < 250 genes, or > 20% mitochondrial genes were excluded. Automated cell classification was performed using *SingleR* with the Human Primary Cell Atlas Dataset serving as reference (Aran et al., 2019). To better define the malignant plasma cell population, we used *inferCNV* to call copy number variants (CNV) in B cells using non-B cells as the germline reference (https://github.com/broadinstitute/inferCNV). The average expression per sample was computed using the *AverageExpression* function in Seurat. We conducted a differential expression analysis comparing responders and non-responders. Subsequently, we applied Gene Set Enrichment Analysis (GSEA) using the *fgsea* R package, utilizing the same gene sets as previously described. Genes were ranked based on the statistics obtained from the differential expression analysis with the Wilcoxon test, and we applied p-value corrections for multiple testing using the Benjamini-Hochberg false discovery rate method.

### Flow cytometry

PB and BM samples were collected in heparinized tubes at baseline and at specified time points during treatment [baseline, at follow up every 3 months during therapy (PB only), response to therapy and progression disease (PD)]. White Blood Cells (WBC) obtained after red blood cell lysis were evaluated using real-time flow cytometry or frozen in a solution of FBS containing 10% DMSO. Immune phenotyping and quantification of T-cells, B-cells, monocytes, Vψ9Vο2 T-cells, NK cells, MDSC, T-reg, and B-reg were performed serially on BM and PB samples. Cells were washed once with a solution containing PBS and 1% FBS before staining. For membrane immunofluorescence, the cells were incubated with their respective anti-human monoclonal antibodies (mAbs) at 4 ° C in the dark for 30 minutes. Afterward, the cells were washed twice and fixed with para-formaldehyde-PBS 1% before the cytofluorimetric acquisition. Cell surface and intracellular proteins were targeted with fluorescinated isocyanine (FITC), r-phycoerythrin (PE), chlorophyll protein peridine (Per-CP) or allophycocyanin (APC), conjugated with mAb, according to the list in **Supplemental Table 13**. For FOXP-3 intracellular staining, cells were fixed and permeabilized using the FoxP3 Staining Buffer Set according manufacturer instructions (Miltenyi #130-093-142). Data were acquired with FACS Calibur cytometers and analyzed using FlowJo software as previously reported. Minimal residual disease was measured by Next generation flow cytometry (NGF), according to Euroflow guidelines (Flores-Montero et al., 2017),

### Statistical analysis

The comparison tests of the immune-cell types in different groups at different time points was performed with Wilcoxon test two sided. All p-values are two sided if not specified otherwise. To assess the concordance between BM and PB immune cell composition, we performed a correlation analysis (Spearman coefficient) between any BM immune feature and its PB counterpart. Association of categorical variables with EFS were performed in a univariable fashion, using Kaplan-Meier curves and a log-rank test. EFS was measured from the date of start of treatment to the date of progression. Deaths from causes other than progression were censored.

### Ethics and data accessibility

These data have been submitted to EGA under accession numbers EGAD00001010161 (whole genome sequencing). The Kydar scRNA-seq data are publicly available at the National Center for Biotechnology Information’s Gene Expression Omnibus (accession no. GSE161195). The CoMMpass data was downloaded from the MMRF researcher gateway portal (https://research.themmrf.org). CoMMpass raw data are accessible on dbgap: phs000748.v1.p1.

## AUTHOR CONTRIBUTIONS

FM designed and supervised the study, analyzed the data, and wrote the paper.

AL, MD designed and supervised the study, collected the samples, and wrote the paper. MM supervised the study, analyzed the data, and wrote the paper.

BZ, CG, analyzed the data and wrote the paper.

GB and ES collected the samples.

DC, BD, MP, OL, EG: analyzed the data.

BB, NB, PC, SO, and MB designed the study and collected the sample.

## Data Availability

These data (whole genome sequencing) will be submitted to EGA. The Kydar scRNA-seq data are publicly available at the National Center for Biotechnology Information Gene Expression Omnibus (accession no. GSE161195). The CoMMpass data was downloaded from the MMRF researcher gateway portal (https://research.themmrf.org). CoMMpass raw data are accessible on dbgap: phs000748.v1.p1.

## ACKNOWLEDGMENTS

This study was funded and supported by Associazione Italiana per la Ricerca sul Cancro (AIRC IG 20541).

This work was supported by the Myeloma Solutions Fund (MSF), Paula and Rodger Riney Multiple Myeloma Research Program Fund, the Tow Foundation, Sylvester Comprehensive Cancer Center NCI Core Grant (P30 CA 240139).

FM is supported by the American Society of Hematology (ASH), Leukemia & Lymphoma Society (LLS), and by International Myeloma Society (IMS).

N.B. Is supported by the Associazione Italiana per la Ricerca sul Cancro (AIRC Investigator Grant n. 25739) and by the European Research Council under the European Union’s Horizon 2020 research and innovation program (grant agreement No. 817997)

