## Supplementary Figures for "Genomic and immune determinants of resistance to anti-CD38 monoclonal antibody-based therapy in relapsed refractory multiple myeloma"

### SUPPLEMENTAL FIGURE LEGEND

**Supplemental Figure 1. Genomic landscape of 28 patients treated with Dara-Rd. a)** Heatmap showing the 31 MM driver mutated genes among 28 patients treated with Dara-Rd. Two additional patients treated with other Dara-based regimens were included as well. **b)** The cumulative copy-number plot (top) and cumulative structural variant plot (bottom) across the entire series.

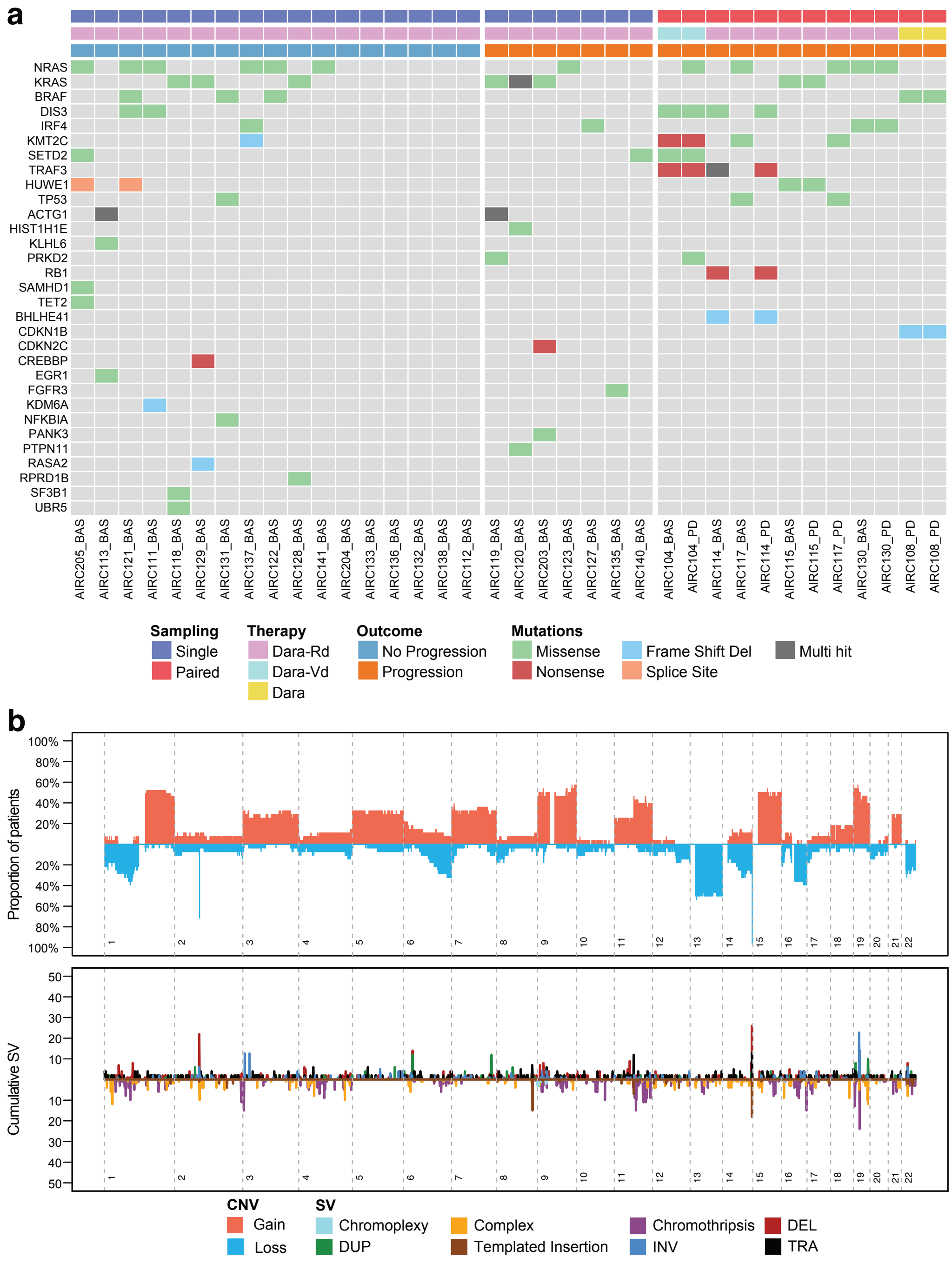

**Supplemental Figure 2. Gene Set Enrichment Analysis (GSEA) for two aneuploidies associated with outcome, using the CoMMpass public dataset.** Heatmap showing the 52-hallmark gene-sets tested in this analysis. The X and * indicate FDR<0.05 and FDR<0.1 respectively, while the bold pathways are significant in both aneuploidies and in the Kydar single cell RNA data set. NES: normalized enrichment score.

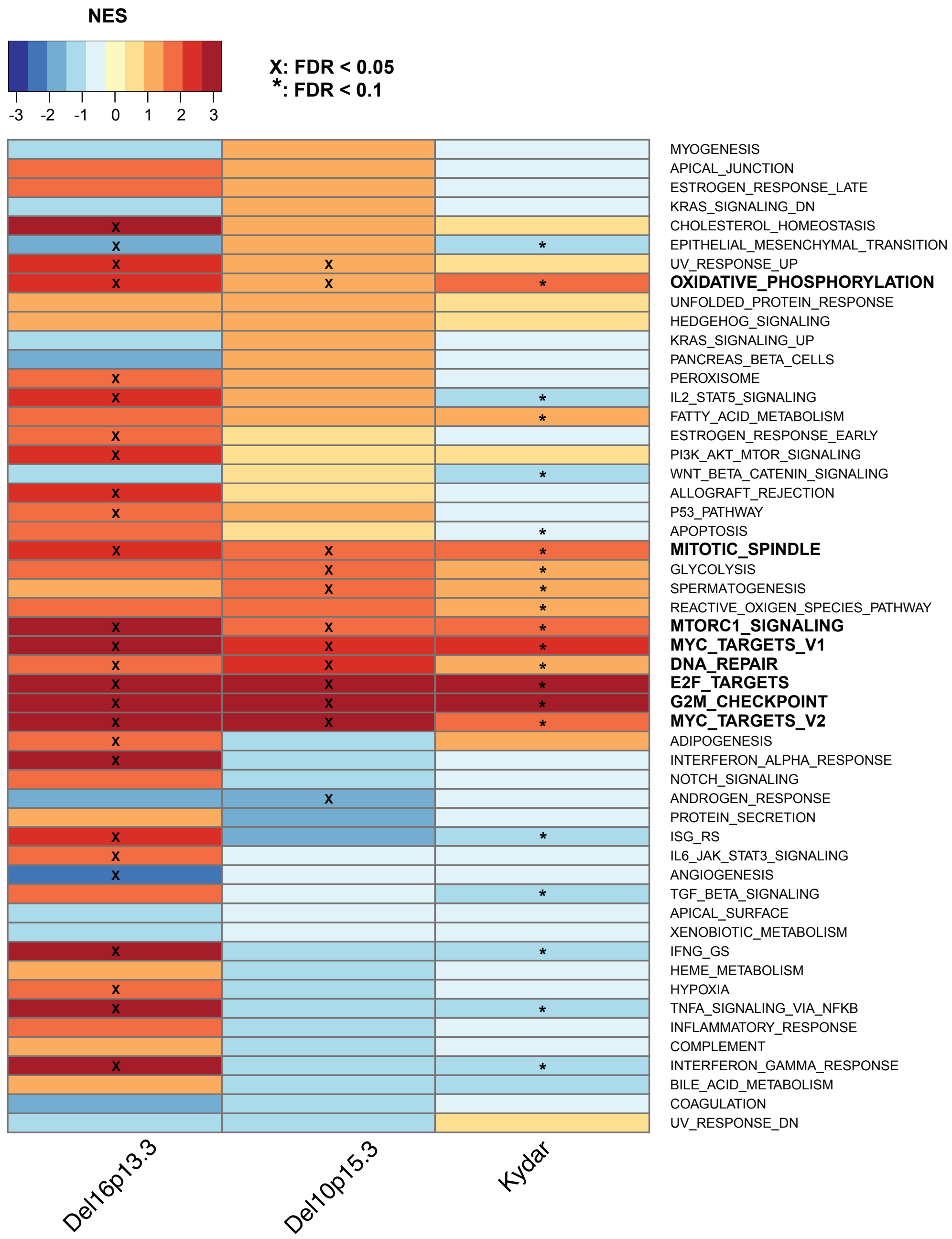

**Supplemental Figure 3. Correlation analysis between BM and PB immune composition at baseline. a)** Heatmap showing the Spearman coefficient per each contrast. The color scale is between 0.5 and 1 (red) and -0.5 and -1 (blue). **b)** Scatterplot with the linear regression line (red) between all the number of cells for each immune population in BM and PB at baseline. **c)** Scatterplot using relative values.

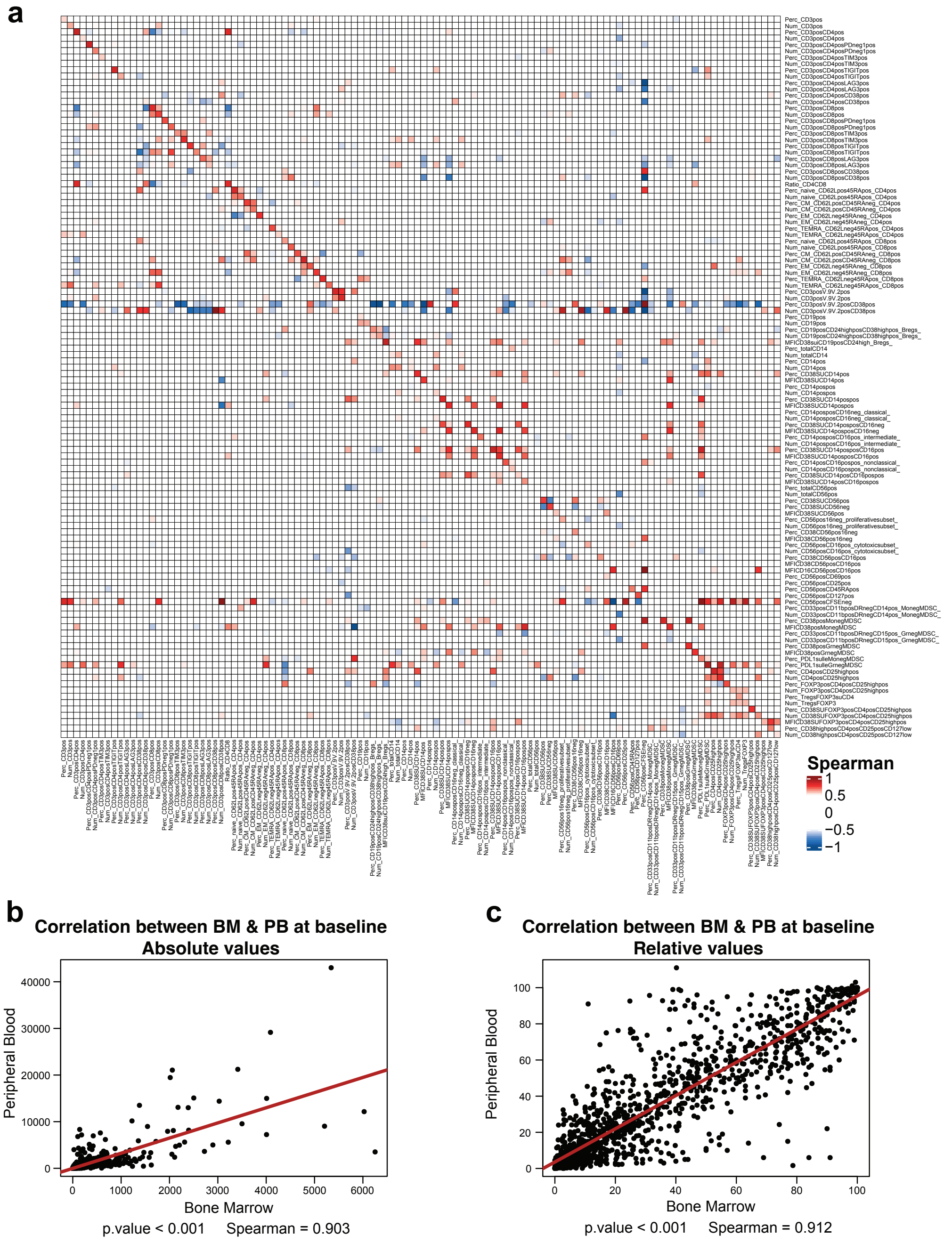

**Supplemental Figure 4. Immune composition of RRMM at baseline. a)** Barplot showing the percentage of CD38 pos for each immune cell population. **b)** Barplots showing the percentage of CD38 pos Gr-MDSC and Mo-MDSC cells in BM and PB samples. **c)** The percentage of Gr-MDSC and Mo-MDSC cells in BM and PB samples at baseline. **d)** Barplots presenting the percentages of CD38 pos monocyte subgroups. **e)** The percentage of Monocyte cells at baseline in both BM and PB samples. **f)** Barplot showcasing the percentage of CD38 pos NK cell at baseline in both BM and PB samples. **g)** The percentage of different NK subgroup cells at baseline in both BM and PB samples. The p-values were estimated using Wilcoxon test two sided; *≤0.05, **≤0.01, ***≤0.001, ****≤0.0001. NonCl: non classical; Int: Intermediate.

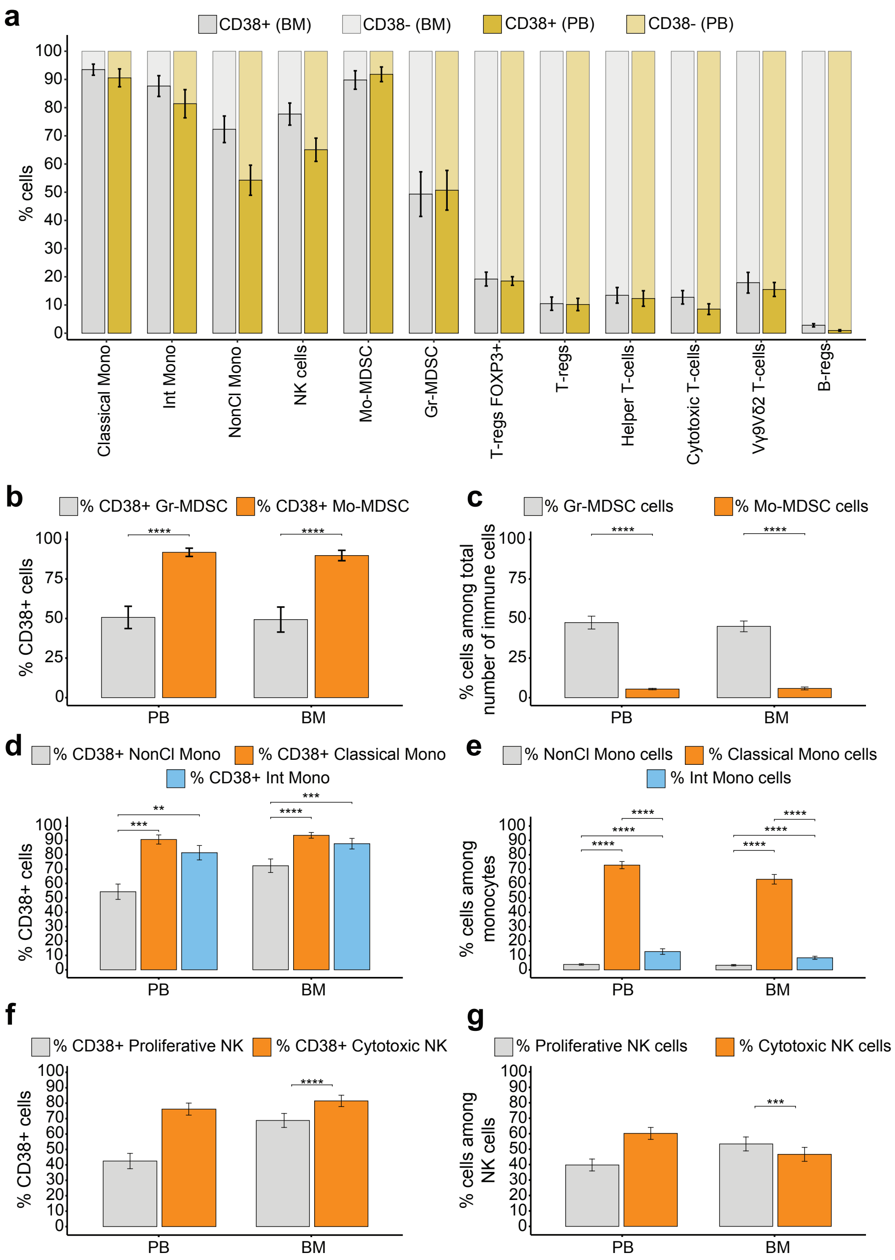

**Supplemental Figure 5. Association between immune composition at baseline with outcome. a-b)** The percentage of CD38 pos immune cell in durable responders and progressors in BM (**a**) and in PB (**b**) samples. **c-l)** Boxplots showing the associations between immune cells at baseline in BM and PB samples and outcome. All p-values were estimated using Wilcoxon test two sided.

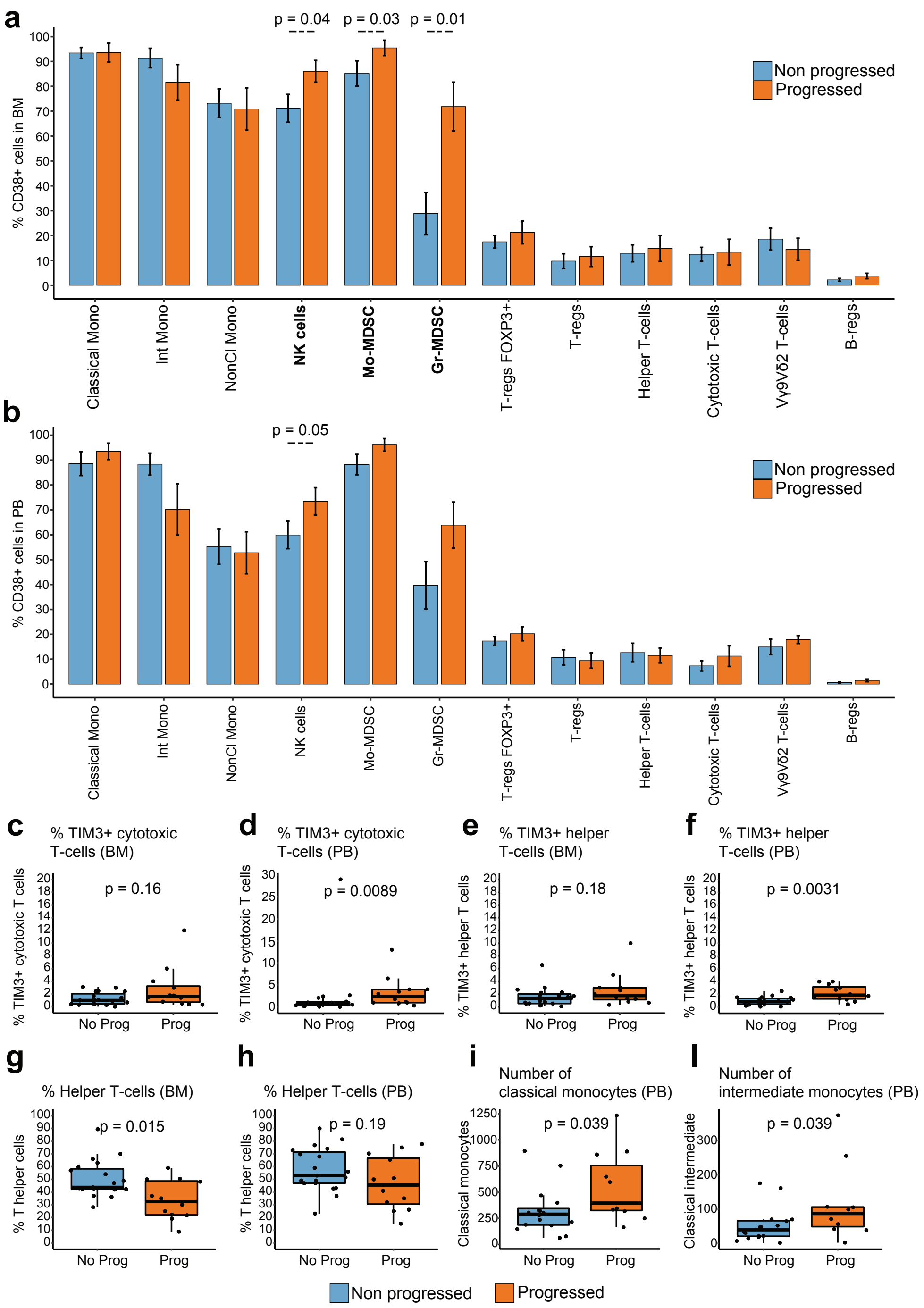

**Supplemental Figure 6. Immunomodulation overtime. a)** Bubble chart about the percentage of CD38 pos immune cell in BM samples before and after Dara-Rd in patients who achieved a durable response. **b)** Boxplots showing the percentage of CD38 pos intermediate monocyte overtime. **c-d)** Boxplots showing the absolute number of intermediate and non-classical monocytes overtime and based on response to Dara-Rd treatment (durable responders are in blue and progressors in orange). **e)** Boxplots showing the number of CD38 pos CD4+ CD25 CD127low T-reg cells overtime. **f-n)** Boxplots showing the immune cells associated with the significant genomic drivers. All p-values were estimated using Wilcoxon test two sided. The entire list of p-values is available in the **Supplemental Table 9-10**.

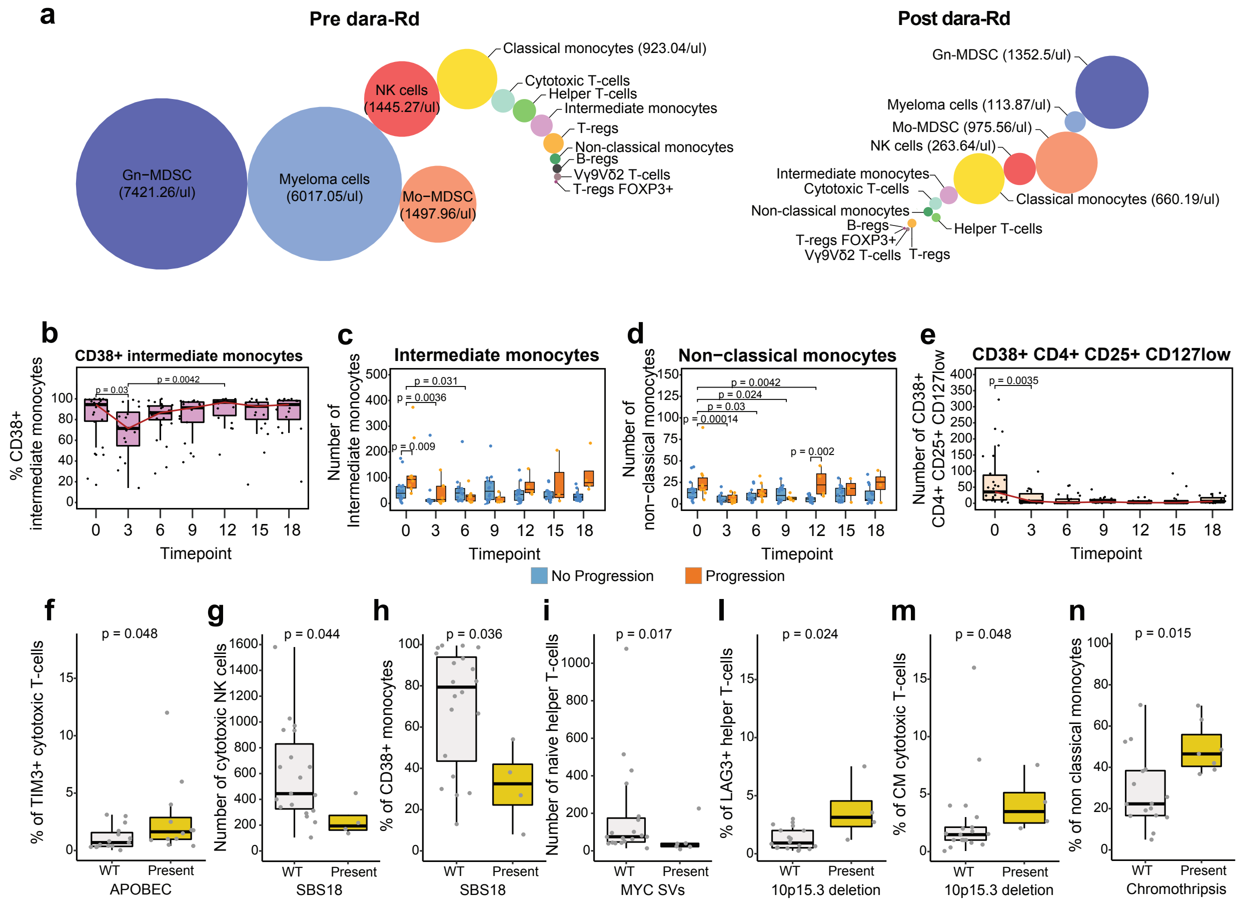
